# Multiomics machine learning identifies inflammation molecular pathways in prodromal Alzheimer’s Disease

**DOI:** 10.1101/2023.03.02.23286674

**Authors:** Alicia Gómez-Pascual, Talel Naccache, Jin Xu, Kourosh Hooshmand, Asger Wretlind, Martina Gabrielli, Marta Tiffany Lombardo, Liu Shi, Noel J. Buckley, Betty M. Tijms, Stephanie J. B. Vos, Mara ten Kate, Sebastiaan Engelborghs, Kristel Sleegers, Giovanni B. Frisoni, Anders Wallin, Alberto Lleó, Julius Popp, Pablo Martinez-Lage, Johannes Streffer, Frederik Barkhof, Henrik Zetterberg, Pieter Jelle Visser, Simon Lovestone, Lars Bertram, Alejo J. Nevado-Holgado, Alice Gualerzi, Silvia Picciolini, Petroula Proitsi, Claudia Verderio, Juan A. Botía, Cristina Legido-Quigley

## Abstract

Mild Cognitive Impairment (MCI) is a phase that can precede Alzheimer’s Disease (AD). To better understand the molecular mechanisms underlying conversion from MCI to AD, we applied a battery of machine learning algorithms on 800 samples from the EMIF-AD MBD study. The cohort comprised participants diagnosed as 230 normal cognition (NC), 386 MCI (with longitudinal data on AD conversion or remaining stable) and 184 AD-type dementia. Data consisted of metabolites (n=540) and proteins (n=3630) measured in plasma coupled to clinical data (n=26). Multiclass models selected oleamide, MMSE and the priority language as the most confident features while MCI conversion models selected pTau, tTau and JPH3, CFP, SNCA and PI15 proteins. These proteins selected for MCI conversion have been previously associated with AD-related phenotype. Oleamide, a possible anti-inflammatory, prompted in-vitro experiments in rodent microglia. The results demonstrated that disease-associated microglia synthesize oleamide which were excreted in vesicles. In addition, plasma vesicles extracted from participants with AD showed elevated oleamide levels compared to controls (P<0.05). This study uncovered MCI conversion pathways that involve inflammation, neuronal regulation and protein degradation.

**Graphical abstract:** 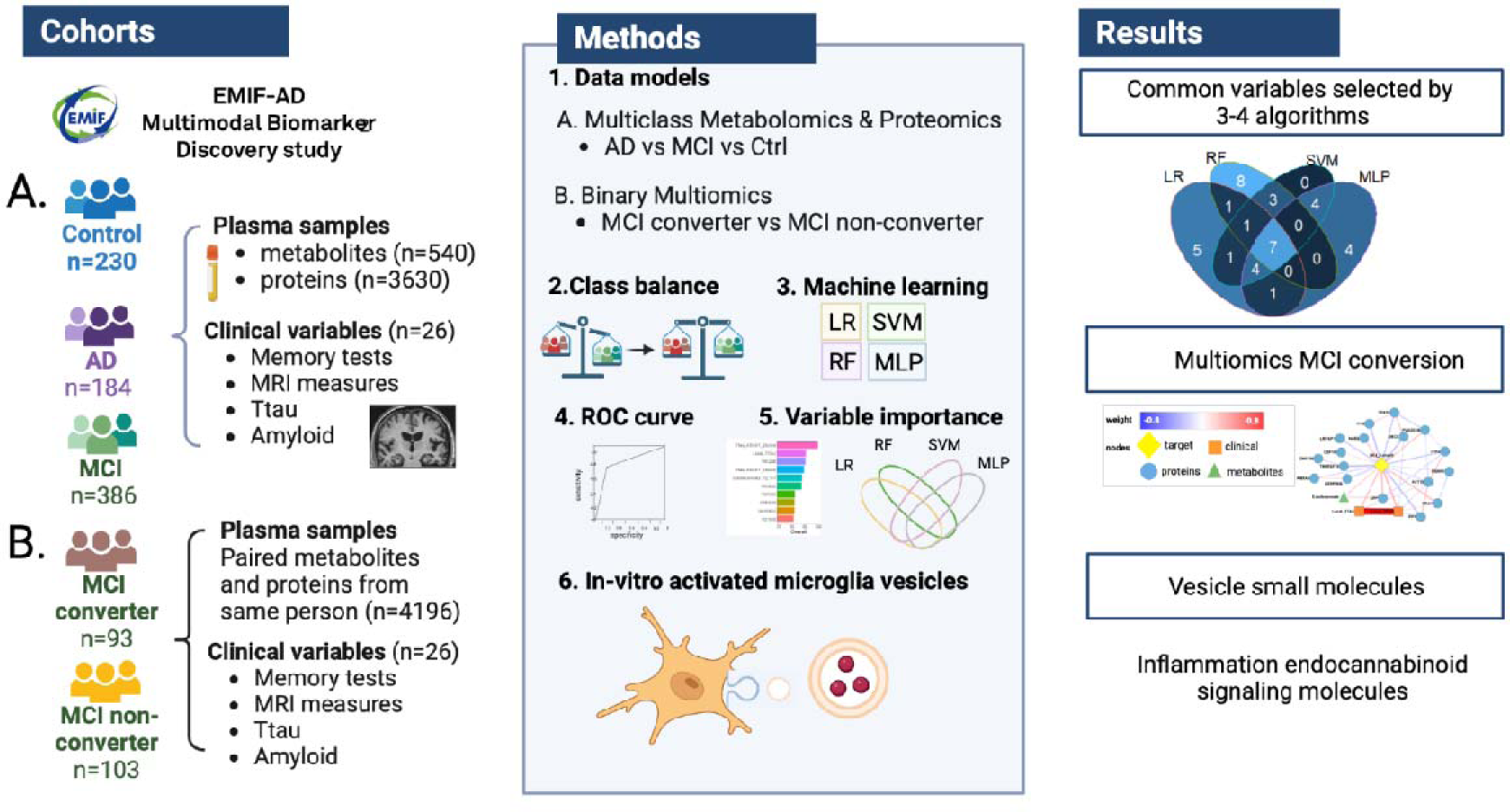

## Introduction

Mild cognitive impairment (MCI) is defined as the symptomatic predementia stage characterized by objective impairment in cognition that does not interfere notably with activities of daily life^1^. It is estimated that over 15% of community dwellers have MCI. The prevalence of MCI increases with age and decreases with education, and it is a heterogeneous and unstable condition^2^. MCI individuals who eventually progress to Alzheimer’s Disease (AD) diagnosis are classified as MCI converters (cMCI), while those who remain stable or improve are classified as MCI stable (sMCI). It has been reported that approximately 29% of individuals with prevalent or incident MCI will progress to dementia, while 38% will revert back to a normal cognition diagnosis^3^.

The conversion of MCI to dementia has been studied using various data sources, including neuropsychological tests, demographic information, neuroimaging (both structural and functional), genetics, and cerebrospinal fluid biomarkers, either alone or in combination^4–10^. With the advancement of high-throughput sequencing technologies, there has been increased interest in “omics” data, which measure different molecules such as the genome, transcriptome, proteome, metabolome, and lipidome^11^. The most common method to quantify proteins, metabolites and lipids is mass spectrometry, which measures the mass-to-charge ratio of ionized molecules^12^. These measures are taken from biofluids such as plasma and cerebrospinal fluid, as structural and functional changes in the brain can be reflected in these fluids^13^. Many studies investigating MCI progression to AD use traditional methods such as basic statistical tests, correlation and similar metrics^10^ as well as models such as Cox regression^5,10^ or Bayesian networks^9^. However, the high dimensionality of omics data is particularly suited for machine learning (ML), a computational approach that delivers further insight into complex data^14^. Previous research using omics, imaging and clinical data to study AD applied ML algorithms such as lasso regression^4,9^, support vector machines (SVM)^4^ as well as different deep learning algorithms such as multiple kernel learning^4,6^ and multimodal recurrent neural network^8^. In this same line, in our previous works, we employed random forest, SVM and regression-based approaches for the study of omics related to AD^15–17^.

In order to identify early disease pathways involved in the conversion to AD, we analyzed both proteomics (n=3630) and metabolomics (n=540) data from 800 participants from the European Medical Information Framework for Alzheimer’s Disease Multimodal Biomarker Discovery Study (EMIF-AD MBD)^18^. The cohort comprised 230 donors diagnosed with normal cognition (NC), 386 MCI participants and 184 AD-type dementia participants. Our study aims to address two main research questions: 1) which proteins and metabolites are most relevant in differentiating between NC, MCI, and AD? and 2) which proteins and metabolites are most relevant in differentiating between sMCI and cMCI? To specifically examine MCI progression to AD, we created a smaller cohort with same person paired proteomics and metabolomics data from 103 sMCI and 93 cMCI participants. We applied four ML algorithms to create both multiclass models and MCI conversion models. Multiclass models selected oleamide (an endocannabinoid), MMSE and the priority language as the most confident features while MCI conversion models selected pTau, tTau and JPH3, CFP, SNCA and PI15 proteins. Since another endocannabinoid (EC), anandamide, was previously found in microglia cells^19^, we investigated oleamide in the same cell type. In addition, we identified properdin (CFP), a plasma glycoprotein that activates the complement system of the innate immune system, as a relevant molecule involved in MCI conversion, which motivated a focus on neuroinflammation. Our results in people with an MCI conversion diagnosis indicate the presence of pathways related to inflammation, sedation, neural and protein degradation.

## Results

### Multiclass classifiers for NC, MCI and AD-type individuals

Multiclass models were created to classify NC, MCI and AD donors using four different machine learning algorithms in two different datasets: proteins with clinical covariates and metabolites with the same covariates. For both approaches, a similar performance was observed between the four different algorithms in the training step, with a mean accuracy of 0.86 for the proteins and 0.826 for metabolites (Table 1, Supplementary Figure 1). However, when evaluating the models on the test set, SVM performance with both proteins and metabolites was superior. Within the protein models, the mean accuracy of all algorithms on the test set was 0.717 while SVM obtained a 0.882 of accuracy. Analogously, when dealing with the metabolites, the overall accuracy on the test set was 0.717 but SVM obtained an accuracy of 0.854 (Table 1). A possible explanation was that SVM was less prone to overfitting for diagnosis prediction, therefore, it got a better test accuracy^20^.

**Table 1.** Multiclass models performance for proteins and metabolites separately for each algorithm. Standard errors indicated in brackets represent cross-validation error.

| **Omics data** | **Algorithm** | **Training accuracy** | **Test accuracy** |
| --- | --- | --- | --- |
| Clinical data and proteins | LR | 0.860 (0.034) | 0.735 (0.046) |
|  | SVM | 0.875 (0.025) | 0.882 (0.016) |
|  | RF | 0.854 (0.049) | 0.729 (0.072) |
|  | MLP | 0.857 (0.049) | 0.701 (0.048) |
| Clinical data and metabolomics | LR | 0.826 (0.074) | 0.717 (0.083) |
|  | SVM | 0.843 (0.062) | 0.854 (0.014) |
|  | RF | 0.872 (0.052) | 0.763 (0.075) |
|  | MLP | 0.844 (0.063) | 0.701 (0.070) |

Despite the algorithms perform similarly during training, the approaches largely identified different relevant features. For the protein models, the top 20 most predictive clinical and molecular features were extracted for each algorithm and the features overlap between algorithms were represented with a Venn Diagram (Figure 1A, Supplementary Table 1). The four approaches identified different features, and this is because 1) we used algorithms from different families, that is, they use different strategies to identify patterns in the data; 2) this was a high-dimensionality problem, the presence of collinearity (features highly correlated) can also influence feature selection, therefore, we can find complementary but different markers selected by each algorithm. We decided to keep the features selected as relevant by most of the algorithms since we considered they would be more universal (not exclusive to a particular algorithm), hence a candidate to interpret biologically with more confidence.

**Figure 1.**
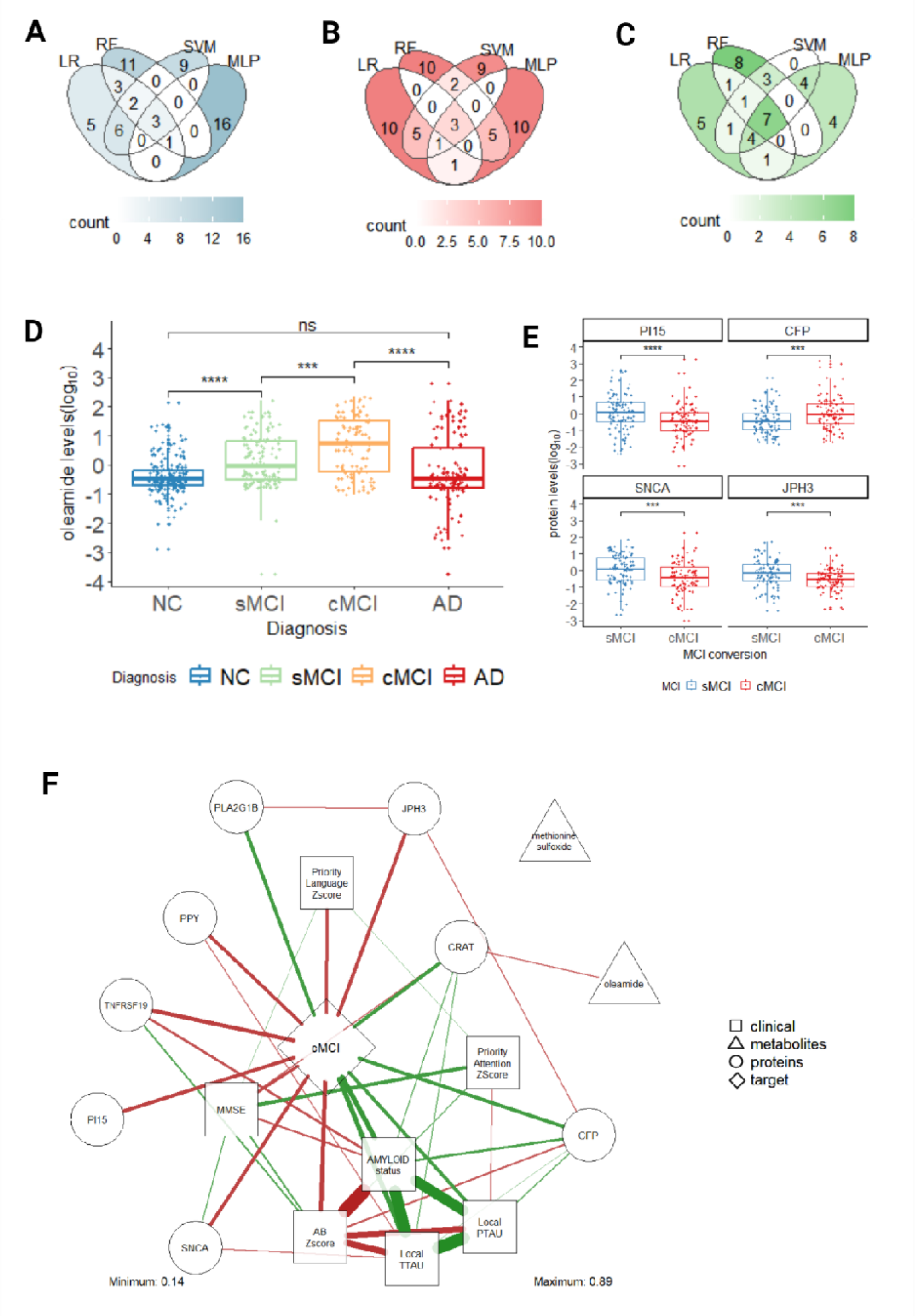
Most relevant clinical features, proteins and metabolites extracted with multiclass models and MCI conversion models. Venn diagram shows the overlap of the top 20 most relevant predictors of each algorithm for **(A)** multiclass models of proteins, **(B)** multiclass models of metabolites and **(C)** MCI conversion models of proteins and metabolites; **(D)** Oleamide level distribution is represented for NC (n=203), sMCI (n=128), cMCI (n=99) and AD (n=136) donors. Differences in oleamide levels between pairs of groups were estimated using a Wilcoxon test. **(E)** Main proteins levels distribution is represented for sMCI (n=291) and cMCI (n=100) participants. Differences in the protein levels between sMCI and cMCI were estimated using a Wilcoxon test. **(F)** Correlation network including the variables selected as relevant by at least three of the four algorithms proposed in all the approaches: multiclass models for proteins, multiclass models for metabolites and MCI conversion models. Pearson correlations were estimated using the sMCI (n=103) and cMCI (n=91) donors with paired data (proteomics and metabolomics). Only significant correlations were shown (*P*<0.05). The minimum and maximum strength of the correlation is shown in the figure. Positive correlations were represented with green color, negative correlations with red. The color saturation and the width of the edges corresponds to the absolute weight and scale relative to the strongest weight in the graph. Features were grouped into target, clinical, proteins or metabolites. ns, non-significant; *, P<0.05; **, P<0.01; ***, P<0.001; ****, P<0.0001.

In this regard, three clinical variables were selected as relevant by all the algorithms, these were: priority attention z score, priority language z-score and priority memory delayed z score. Amyloid status and MMSE were selected by all algorithms except one. Five proteins were selected in only two algorithms (none in 3 and 4), these were: Apolipoprotein D (Apo D), Trypsin-1 (PRSS1), Procollagen C-endopeptidase enhancer 1 (PCOLCE), B melanoma antigen 3 (BAGE3) and Semaphorin-6C (SEMA6C).

The same procedure was repeated with metabolite models. Two clinical variables, MMSE and priority language z-score, were selected by all the algorithms (Figure 1B, Supplementary Table 1). In addition, one metabolite - oleamide – was selected by all four algorithms. Pairwise comparisons of oleamide levels were computed between the four groups of participants (NC, sMCI, cMCI and AD) (Figure 1D). Oleamide levels were increased in sMCI donors compared to NC (Wilcoxon test *P*<8.721 · 10^−10^, W = 7793). Oleamide levels increased even more in cMCI donors compared to sMCI (Wilcoxon test *P*<2.696 · 10^−4^, W = 4548). However, oleamide levels decreased in AD donors compared to cMCI donors (Wilcoxon test *P*<1.593 · 10^−8^, W = 9640.5). Finally, AD donors reached similar oleamide levels compared to NC donors.

Changes in oleamide levels were significantly associated with diagnosis status after adjusting for age and sex covariates including all donors (ANCOVA *P*<2 · 10^−16^, F=28.095, df=3) and only MCI donors (ANCOVA *P*<4.3 · 10^−4^, F=12.779, df=1). These same models shown that sex had no influence on oleamide levels while age had a significant influence on oleamide levels when including all donors (ANCOVA *P*<1.22 · 10^−8^, F=33.452, df=1) and only MCI donors (ANCOVA *P*<7.72 · 10^−6^, F=20.979, df=1).

Additionally, methionine sulfoxide was selected by three algorithms. AB Zscore, glycerophosphorylcholine (GPC), glycosyl-N-tricosanoyl-sphingadienine-d18:2/23:0, iminodiacetate (IDA), N1-methylinosine, serotonin, 3-hydroxyhippurate and ximenoylcarnitine (C26:1) were selected by two algorithms. The features selected as relevant by each algorithm were reported in Supplementary Table 1.

### Classifier proteins involved in MCI conversion

MCI conversion models reported a mean ROC of 0.64 across the different algorithms, with a mean sensitivity of 0.614 and mean specificity of 0.582 (see Supplementary Table 2). All algorithms performed similarly in terms of metrics. The overlap of the top 20 most relevant variables of the four models was represented with a Venn Diagram (Figure 1C, Supplementary Table 1). Seven features were selected as relevant in all models. Among them, three clinical variables, local pTau and tTau, tTau Zscore were selected. In addition, four proteins were selected by all the algorithms: Peptidase inhibitor 15 (PI15), Properdin (CFP), Alpha-synuclein (SNCA) and Junctophilin-3 (JPH3). Proteins selected by three algorithms were Pancreatic hormone (PPY), Phospholipase A2 (PLA2G1B), Carnitine O-acetyltransferase (CRAT) and Tumor necrosis factor receptor superfamily member 19 (TNFRSF19). Finally, for two algorithms, 5-dodecenoic acid (12:1(n-7)) metabolite was selected as relevant. None of these clinical features, proteins or metabolites was also selected as relevant by the multiclass models. All proteins selected as relevant for MCI conversion, except PLA2G1B, were previously associated with AD-related phenotype. Most of these proteins were found to have evidence for brain eQTL and RNA expression change in the AD brain (Supplementary Table 3).

CFP was increased in cMCI compared to sMCI (Wilcoxon test *P*<6.99 · 10^−4^, W = 6010), while JPH3 (Wilcoxon test *P*<6.072 · 10^−4^, W = 3348), PI15 (Wilcoxon test *P*<9.625 · 10^−5^, W = 3164) and SNCA were decreased in cMCI compared to sMCI (Wilcoxon test *P*<1.672 · 10^−4^, W = 3217) (Figure 1E). After adjusting for age and sex covariates, we found significant differences between cMCI and sMCI participants for PI15 protein (ANCOVA *P*<5.96 · 10^−4^, F= 12.194, df=1), SNCA protein (ANCOVA *P*<7.08 · 10^−4^, F= 11.853, df=1), JPH3 protein (ANCOVA *P*<9.91 · 10^−4^, F= 11.191, df=1) and CFP protein (ANCOVA *P*<6.39 · 10^−4^, F= 12.057, df=1). We found that sex had no influence on the levels of these proteins for MCI participants while age only had a significant effect on SNCA protein levels (ANCOVA *P*<0.0305, F= 4.749, df=1).

The features selected by three of the four algorithms from multiclass models and MCI conversion models were represented in a correlation network to visualise any additional relationships between the selected molecules. MCI conversion was the target outcome and proteins or metabolites were linked to each other (Figure 1F). The network shows the strength of the relationships as Pearson correlations. Only significant correlations were shown in the figure (*P*<0.05). All the protein molecules correlated with cMCI (corr > |0.3|), however, the metabolites had low correlation (corr< |0.1|). A maximum correlation |0.97| was observed between amyloid z-score and status measures. The results shown that cMCI had relatively higher levels of brain biomarkers such as tau and amyloid compared to sMCI while MMSE score was lower for cMCI individuals compared to sMCI individuals.

To investigate the prediction ability of these selected clinical features, proteins and metabolites, we employed the same four ML algorithms to predict MCI conversion with a nested cross-validation. The final models showed a mean accuracy of 0.745 (0.015), with a very similar performance between the different algorithms (Supplementary Table 4).

### Oleamide in in-vitro disease-associated microglia

We conducted oleamide experiments in microglia culture from rodents and humans motivated by anadamide’s potential metabolism within microglia and their vesicles based on previous studies^19,21,22^. The LC-MS quantitation for oleamide was carried out on microglia cultures and their secreted extracellular vesicles (EVs). The concentration of oleamide was normalized by the total proteins in cells and EVs. Details about the LC-MS quantification method are available in Methods and Supplementary Material sections.

The amount of oleamide was higher in microglia compared to the supernatant and further concentrated in EVs (n=3) (Figure 2A). Moreover, it can be observed that activated microglia contained higher amounts of oleamide compared to unstimulated cells. We also observed a similar amount of oleamide in mouse and rat microglial EVs (two independent experiments, three measurements) (Figure 2B).

**Figure 2.**
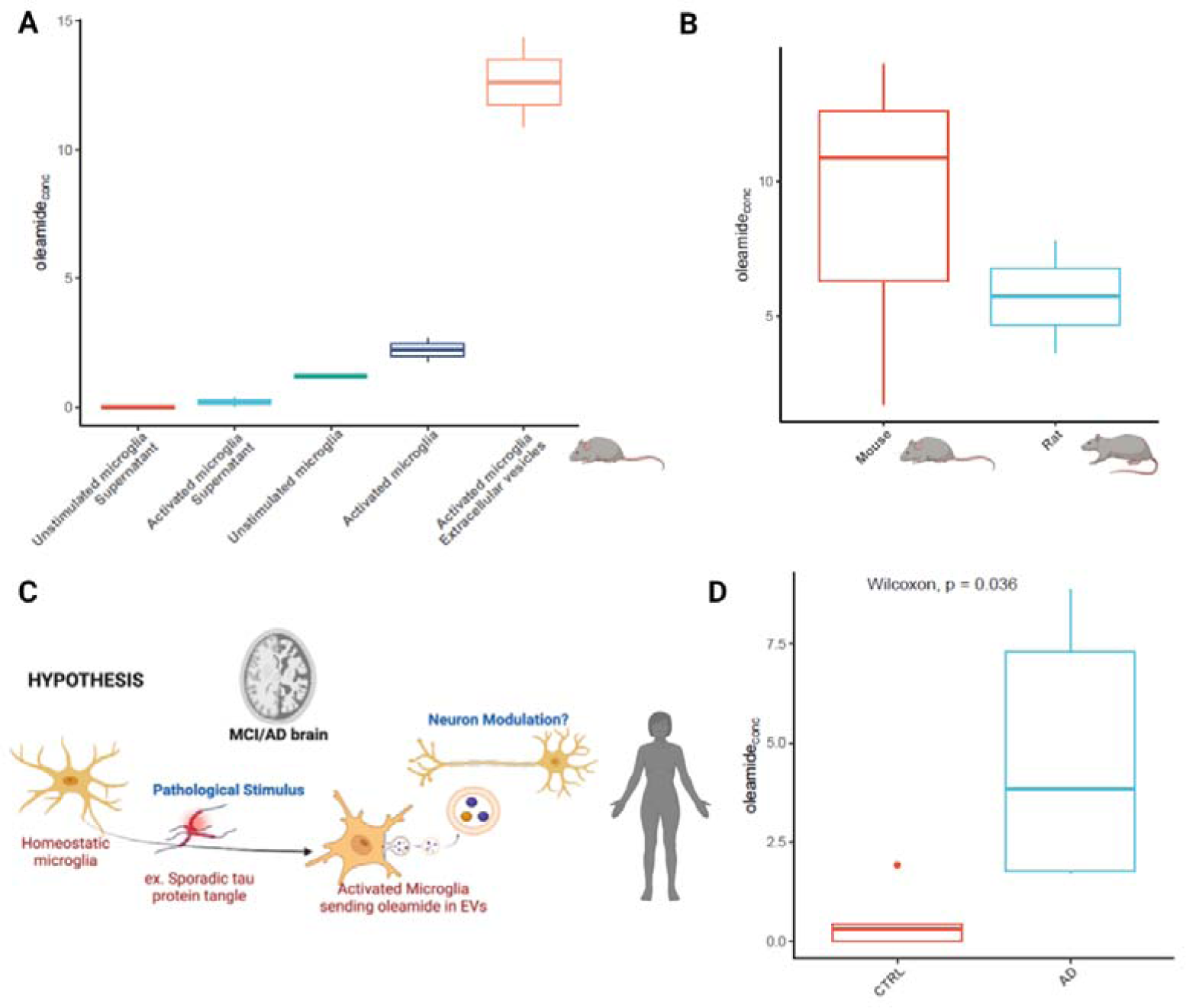
Oleamide in microglia and vesicles. **(A)** Oleamide concentration in pure murine primary microglia per state and EVs. Concentration is normalized by protein amount in cells and EVs (n=2 independent experiments). **(B)** Oleamide in rodent microglia EVs. Oleamide concentration is normalized per protein amount in EVs isolated from mice (n=3 independent experiments) and rats (n=2 independent experiments). **(C)** Oleamide possible mechanisms. Oleamide is released in EVs from activated microglia, these EVs might modulate or inhibit neuronal transmission via CB1 receptors. **(D)** Concentration of oleamide in EVs isolated from plasma of AD and healthy subjects.

Quantitation for oleamide was carried out on EVs extracted from plasma of five AD and five control subjects. The concentration of oleamide was significantly increased in AD (Wilcoxon test *P*<0.05) (Figure 1D). Figure 2C presents a hypothetical mechanism where microglia activated by fibrils in MCI/AD brains release oleamide in EVs, oleamide as an EC could possibly modulate or inhibit neuronal transmission.

## Discussion

The use of artificial intelligence in uncovering disease-specific pathways has gained great interest in recent years. Machine learning models, by their nature, have the ability to analyze big data and potentially detect complex molecular interactions, such as non-linear relationships. In this study, we used the EMIF-AD MBD dataset and applied ML algorithms to identify key molecules involved in the conversion of MCI to AD. Four different algorithms (logistic regression, support vector machines, random forest, and multi-layer perceptron) were used to classify: 1) NC vs MCI vs AD, and 2) cMCI vs sMCI. Further molecular exploration focused on molecules that were consistently identified in four algorithms and thus assumed to be universally linked to disease status.

### Models performance and clinical covariates

The four algorithms showed a similar performance in the training of multiclass models for both proteins (0.86 mean accuracy) and metabolites (0.826 mean accuracy). All the algorithms showed a lower accuracy of prediction for the MCI class, possibly due to the heterogeneity covered by the MCI diagnosis (Supplementary Figure 1). SVM outperformed all other algorithms in the test data, reaching values of accuracy of 0.882 for proteins and 0.854 for metabolites.

The model’s performance was in line with previous work. Stamate et al.^16^ analyzed a previous cohort with fewer individuals (115 AD donors and 242 NC donors) with different ML algorithms to demonstrate that plasma metabolites have the potential to match established AD CSF biomarkers. In this previous work, deep learning model produced an AUC value of 0.85, Extreme Gradient Boosting model reported a 0.88 whereas the random forest model resulted in a 0.85 AUC value. Additionally, a proteomics study by Shi et al.^23^, where ML algorithms were used to classify amyloid positive and amyloid negative participants. Lasso Regression and SVM models parsed a predictive panel composed of 44 proteins, age and the risk gene APOE4 achieving an AUC of 0.68 in a replication group.

Clinical covariates were included in all the models to interpret pathway biology. For example, in the multiclass models, the MMSE test and the priority language score were ranked high for both proteins and metabolites models while these variables did not play an important role in MCI conversion. Instead, Tau-related measures (pTau and tTau) were shown by the binary model to be most relevant. This is interesting because Tau or amyloid measures are deemed to be a more accurate and objective measure for diagnosis. Other important features among the clinical metadata were known risk factors of AD, such as sex, age, years of education and the volume of the hippocampus which is known to be related to the memory function in the brain. However, these were not chosen consistently by the models (see Supplementary Table 1).

### Multiclass models select oleamide

Multiclass models (NC vs. MCI (all) vs. AD) for metabolite models showed that one lipid, oleamide, was selected as a predictor by all algorithms while methionine sulphate was chosen by three. Oleamide is a molecule thought to be synthesized in the brain to aid with sleep^24^ and is a potent endogenous endocannabinoid^25^. The increased levels in the MCI group suggested that sleep deprivation could be involved in memory and cognitive impairment. In a previous study with fewer participants, oleamide, within other ECs, was associated with elevated amyloid levels in the brain^26^. In sensitivity analyses, oleamide was significantly higher in cMCI (see Figure 1D) and it also associated with age. Xie et al.^27^ showed that sleep helps with toxic clearance in the brain and hence, we hypothesised that if oleamide aids with sleep in early AD, this could be an early coping mechanism.

Furthermore, methionine is a sulfur-containing essential amino acid that, among many different functions, it intervenes in the biosynthesis of glutathione to counteract oxidative stress and inflammation^28^. The same proteins were not selected in more than two multiclass models.

### MCI conversion models select mainly proteins

MCI conversion models selected four key proteins: peptidase inhibitor 15 (PI15), properdin (CFP), alpha-synuclein (SNCA), and junctophilin-3 (JPH3). Decreased levels of alpha-synuclein and junctophilin (JPH3) were found in the blood of cMCI participants. Notably, the presence of dual motor and memory symptoms has been shown to increase the risk of developing dementia^42^. The JPH3 protein is particularly interesting because it is a neuron specific protein. JPH3 is regulated in a unique neuron-restricted fashion to control electrical excitability of neurons in different brain regions and is involved in the regulation of intracellular calcium signalling. JPH3 has been linked previously to Huntington-like disease-2^29^. Properdin was the only protein that was elevated in cMCI compared to sMCI. It belongs to the complement system, a well-established pathway of inflammation. This supports the hypothesis that inflammation in the brain exacerbates the progression of AD. The last protein selected by all algorithms was peptidase inhibitor 15 (PI15), which is an inhibitor against trypsin and, although unexplored in brain, may play a role in protein degradation in the central nervous system.

### Correlation network and models with selected molecules

The features selected by three of the four algorithms were represented in a correlation network to visualise links between the selected molecules. At baseline, cMCI donors had relatively higher levels of brain biomarkers such as tau and amyloid and the average MMSE score was lower for cMCI individuals too (mean of 25) compared to sMCI individuals (mean of 26) (demographics supplementary table 5). Two new proteins selected by three ML algorithms, PLA2G1B and CRAT, showed weak positive correlation with conversion. PLA2 is a lipase enzyme involved in the hydrolysis of phospholipids, previously linked to AD^30^. CRAT on the other hand is a carnitine acetyltransferase that has been associated with a severe neurometabolic disorder named Leigh syndrome^31^. The final models with the selected molecules had similar performance with the ML methods with a mean value over seventy.

### Oleamide is detected in microglia and extracellular vesicle (EV)

The decision to conduct oleamide experiments in microglia was motivated by anandamide metabolism within microglia as based on previous studies. Briefly, microglia are known to release a variety of signaling molecules that impact synaptic transmission in response to injury or inflammation and play a crucial role in maintaining balance in neuronal networks^32^. Stella N.^21^ demonstrated that microglia produce in vitro 20-fold higher amounts of ECs than neurons or astrocytes, likely representing the main source of ECs in the inflamed brain. In this regard, Gabrielli, M. et al.^22^ demonstrated that ECs are secreted by microglia through extracellular membrane vesicles and these inhibit presynaptic transmission in target GABAergic neurons. In addition, ECs have been linked to learning, memory and long-term plasticity^33–35^ and can be neuromodulator lipids^36^.

The present study provides the first evidence of oleamide being present also in microglia and enriched in EVs released in the pericellular space. It also showed that EVs from blood contain oleamide in their cargo and this was more abundant in persons with AD. Figure 2C presents a hypothetical drawing where microglia activated by fibrils in MCI/AD brains release oleamide in EVs, oleamide as an EC could neuromodulate or inhibit neuronal transmission.

### Limitations of the study

Machine learning applications to omics data have some limitations, most notably, collinearity, can increase bias, and usually, datasets are still small when compared with imaging studies. Another limitation is that, at baseline, the cMCI and sMCI groups were not matched in CSF biomarkers, as tau and amyloid were elevated in cMCI participants compared to sMCI. Nonetheless, the study has the advantage of having CSF biomarkers and extensive clinical covariates, to help with interpretation.

For the additional studies of selected molecules, the authors were unable to find another existing cohort with both omics data and follow-up outcomes for MCI individuals, which could further validate this study. Moreover, the oleamide measurements were carried out on EVs and microglia in primary culture, an artificial condition far from the *in vivo* setting. In addition, oleamide was measured in EVs from people with AD but in the samples donated by individuals with AD, it was not clear if there were more EVs from brain or more oleamide in the EVs.

In conclusion, our findings revealed inflammation pathways during MCI conversion. The main findings included the lipid oleamide which is linked to sleep and memory and was secreted by microglia via EVs in vitro. A neural protein, JPH3, was also a new potential target, together with PI15 which is a peptidase inhibitor of unknown brain function. The ML based pipeline also confirmed established proteins such as synuclein and protein activators in the complement cascade.

## Methods

### The EMIF-AD Multimodal Biomarker Discovery Study

This study employed data from the European Medical Information Framework for Alzheimer’s Disease Multimodal Biomarker Discovery Study (EMIF-AD MBD)^18^. EMIF-AD MBD is a cross-cohort study consisting of collated data from 11 European cohorts that aims to discover novel diagnostic and prognostic markers for AD-type dementia by performing analyses in multiple biomarker modalities. In the present study, we used data from 230 NC donors, 184 AD participants and 386 participants diagnosed with MCI. Of 386 MCI participants, 100 were later diagnosed with AD-type dementia (defined as AD converting MCI [cMCI]), 219 remained as MCI (defined as stable MCI [sMCI]) and 67 participants do not have this information available. The average follow-up length was 2.5 years. From all cohorts, available data on demographics, clinical information, neuropsychological testing, cognition and Aβ status data were gathered. The cognitive tests used varied across centers and only the Mini Mental State Examination (MMSE)^37^ was administered everywhere and was available for nearly all subjects. At least one test from the following cognitive domains was performed: memory, language, attention, executive functioning and visuoconstruction. For each cognitive domain, a primary test was selected.

Metabolomics data for the current study was acquired by Metabolon Inc. (Morrisville, NC, USA). The relative levels of 883 plasma metabolites were measured in fasting blood samples using three different mass spectrometry methods. Area counts for each metabolite in each sample were extracted from the raw data. The extracted area counts were then normalized to correct for variation resulting from instrument inter-day tuning differences. Metabolite levels below limit of quantification were replaced with 1 while metabolites with more than 20% missing data were excluded for the further analysis. Missing values were imputed with the k-nearest neighbour algorithm. Subsequently, the metabolomics data were log transformed to get a change in scale that is more convenient for this type of analysis and then each metabolite was further scaled to have a mean value of 0 and a standard deviation value of 1. We discarded metabolites with more than 3SD. After data preprocessing, 540 metabolites were selected.

For proteomics analysis, plasma protein levels were assessed in plasma using the SOMAscan assay platform (SomaLogic Inc.). This aptamer-based assay enabled the simultaneous measurement of up to 3630 proteins. Samples were grouped and measured separately. To ensure data consistency across assay runs, 40 subjects were tested in both batches. A detailed description of this process has been previously published^15,38^.

### Choice of algorithms

Our aim was to discover the most relevant clinical characteristics, proteins and metabolites involved in two different tasks: (1) classifying samples into NC, MCI and AD donors and (2) distinguishing between sMCI and cMCI. For each task, we selected four ML algorithms that belong to four different algorithm families. Each family refers to mathematical equations used to discern patterns in the data. Therefore, by using representatives from four families, we improved the confidence of the molecules selected. Logistic regression is one of the most used machine learning algorithms because of its interpretability. It assumes a linear relationship between each explanatory variable and the logit of the response variable. Random Forest, an algorithm combining multiple decision trees for predictions, handles tabular data well and can capture complex relationships. SVM particularly excels in binary classification tasks and perform well with tabular data, finds the optimal hyperplane that best separates classes within a high-dimensional space and can capture non-linear relationships. Neural networks can manage big and complex tabular data sets, potentially it can capture the most complex patterns between molecules, relevant in the context of AD biology.

Once models were built, the top 20 most relevant features were extracted for each model. Model-specific features were selected because each ML method had heightened sensitivity to features or patterns within the data. After this step we selected features repeated in three and four ML methods. This was done because ML approaches have some limitations, they can be prone to overfitting or picking noise incorporating features that lack true relevance. The presence of collinearity, where certain features are highly correlated, can also influence feature selection. Moreover, if the dataset is too small or lacks the variability necessary for describing a diagnosis, it could result in feature variance across models. Therefore, repeated molecules were considered more universal or independent of the algorithm family. We have previously employed this approach of selecting repetitive features to enhance selected molecule confidence^39^.

### Pipeline to create multiclass models of NC, MCI and AD donors

To identify the most relevant proteins and metabolites implicated in the identification of NC, MCI and AD donors, multiclass models were created. Proteins and metabolites were treated separately in their corresponding pipelines and the same clinical covariates were included in both. Multiclass models based on proteins were created using 3630 proteins and 26 clinical covariates from a total of 230 NC donors, 184 AD donors and 386 MCI donors. Multiclass models for metabolites were created using 540 metabolites with identical clinical covariates from a total of 207 NC donors, 136 AD donors and 276 MCI donors. Clinical covariates were included in all the models. Statistics of the clinical characteristics of this dataset have been previously published by Shi et al.^23^

The same pipeline was applied to create both multiclass models for proteins with covariates and the multiclass models for metabolites with covariates. Classes were first balanced using the SMOTE approach^40^ with the ‘imbalanced-learn’ python package. A 5-fold cross-validation repeated three times was applied with the ‘Repeated Stratified K-Fold’ function when fitting the models to address overfitting due to the shape of the data. Four different machine learning algorithms were applied with the ‘Scikit-learn’ python package^41^. The algorithms employed were logistic regression (LogisticRegression), random forest (RandomForestClassifier), support vector machines (SVM) and artificial neural network (MLPClassifier). Each algorithm’s hyperparameters were optimized using this same package. The performance of each algorithm was evaluated using ROC curves (Scikit-learn and matplotlib packages)^42^.

The most relevant variables for each model were extracted using feature importance methods from the Scikit-learn package. For logistic regression models, feature relevance was represented by the magnitude of the coefficient of the model. For random forest models, feature importance was obtained with “featur<u>e_importances</u>_” function, calculated as the mean and standard deviation of accumulation of the impurity decrease within each tree. As SVM and MLP models do not incorporate specific approaches for calculating feature importance, we used a general approach based on predictor permutation through the ‘sklearn.inspection.permutation_importance’ function, where the values in each feature column were shuffled, the effect on the model prediction accuracy was observed and these steps were repeated for all features. Finally, the features selected as relevant (top 20) by at least three of the four algorithms were selected for interpretation.

### Pipeline to create MCI conversion models

To get more specific molecules to MCI conversion to AD, binary models that classify MCI donors into cMCI or sMCI were created using the same ML-based approach. For this purpose, only MCI donors with MCI conversion information and both proteins and metabolites from the same individual were used, taking a total of 103 sMCI and 93 cMCI. The descriptive statistics of clinical characteristics for this dataset are reported in Supplementary Table 4. Paired proteins (n=3630), metabolites (n=540) and clinical variables (n=26) were integrated in the same model.

The pipeline to create the models were based on the functions from the ‘caret’ R package^43^. First, classes were balanced using a downsampling approach (downSample function). Since the number of samples was much smaller than the original set, the complete dataset was used for training. A 3-folds cross-validation repeated 10 times (‘trainControl’ function) was applied to avoid overfitting. In addition, the same four machine learning algorithms were applied, including logistic regression (‘glmnet’ method), random forest (‘rf’ method), support vector machines (‘svmLinear’ method) and artificial neural networks (‘mlpWeightDecay’ method). Hyperparameter tuning was carried out to define our own grid, included in the ‘tuneGrid’ parameter. The top 20 most relevant features of each algorithm were extracted using the ‘varImp’ function. Among them, the features selected as relevant (top 20) in at least three of the four algorithms were selected.

To investigate the prediction ability of the final selected clinical features (see also correlation network molecules), proteins and metabolites, we employed the same four ML algorithms to predict MCI conversion with a nested cross-validation (5 outer folds and 5 inner folds).

### Univariate analysis

Additional univariate analyses were performed. To test the changes in the main proteins and metabolites levels between the four diagnostic groups (NC, sMCI, cMCI and AD), pairwise comparisons were carried out with Wilcoxon test (two-side test) with the wilcox.test function from the stats R package. In addition, we tested differences in proteins or metabolites levels adjusting for age and sex covariates with ANCOVA analysis using ‘aov’ function from the stats R package.

### Correlation network

For the multiclass models and MCI conversion models from each approach, the features selected as relevant by at least three of the four algorithms were included. All these features were put together using a network-based approach, where MCI conversion is the target, and the rest of the features (clinical features, proteins or metabolites) can be linked to the target and between themselves. To this end, a correlation network was created. First, correlations were estimated using the ‘cor_auto’ function from the ‘qgraph’ R package^44^. Then, the correlation network was represented using the ‘qgraph’ function from this same package (graph=“cor”).

### Further studies of selected proteins and oleamide

We investigated whether the proteins selected as relevant were previously associated with AD-related phenotype using two different resources: 1) a systematic review from Kiddle, S. J. et al (2014)^45^, where a list of 21 published discovery or panel-based blood proteomics studies of AD was reviewed and 2) Agora database, a web application that hosts high-dimensional human transcriptomic, proteomic, and metabolomic evidence for whether or not genes are associated with Alzheimer’s disease (https://agora.adknowledgeportal.org/).

### Microglia in-vitro experiments

#### Rodent microglial culture preparation and EV isolation

Pure murine and rat primary microglia, established as in Gabrielli et al^46^. Independent experiments were performed as independent cell preparations and from those several replicates were collected. Microglia have been stimulated with 1:20 Granulocyte-Macrophage Colony-Stimulating Factor (GM-CSF) from murine GM-CSF-transfected X63 cells^47^. GM-CSF is a member of the colony-stimulating factor superfamily that induces microglial proliferation, migration and upregulation of surface markers^48^. Supernatants have been cleared from cell debris before storing. Cells have been scraped in physiological solution, pelleted and stored in methanol. Extracellular vesicles (EVs) have been isolated through differential centrifugation from the cell supernatant upon 30 minutes ATP stimulation at 110,000xg^49^ and stored at −80C°.

#### Isolation of EVs from the plasma of AD patient and control group

Plasma samples were collected from five subjects with a diagnosis of Alzheimer’s disease (75.6±2.7 years, 2/3 males/females ratio) and five healthy controls (65.6±3.7 years, 3/2 males/females ratio). All participants or their representatives provided informed written consent, following the protocol approved by the Ethical Committee of Fondazione Don Carlo Gnocchi, according to the declaration of Helsinki. Inclusion criteria for AD were considered: diagnosis of AD according to NIA-AA criteria; mild dementia stage as documented with a Clinical and Dementia Rating (CDR) scale score between 0.5 and 1; absence of psychiatric or systemic illness. To have a global index of cognitive functioning, all subjects performed cognitive evaluation using the Montreal Cognitive Assessment test (MoCA). Human peripheral blood samples of each subject were collected in EDTA-treated tubes (BD Vacutainer, Becton Dickinson). To isolate the plasma, samples were centrifuged at centrifugation at 1300g x 10 min for removing cells and then at 1800g x 10 min for the depletion of platelets. Plasma aliquots were anonymized, aliquoted and stored at −80°C in the biorepository of the Laboratory of Nanomedicine and Clinical Biophotonics of Fondazione Don Carlo Gnocchi (Milan, Italy) until further use. 500 µl of plasma samples were then thawed and centrifuged at 10,000 g for 10 minutes and then used for EVs isolation by size exclusion chromatography (SEC; qEV, Izon, Christchurch, New Zealand), following manufacturer’s instructions. Eluted fractions from 6 to 8 containing EVs in PBS were retained, added with 0.1% DMSO and stored at −20°C until further analysis.

#### Sample preparation

Oleamide was extracted by the addition of 1 ml of ethanol to the dried microglia and extracellular vesicles. The tubes were then vortexed, for 5s, shaken at 1500 RPM for 5 min at 4 °C, vortexed, for 5s, and subsequently centrifuged at 12,000 g for 5 min at 4°C. Next, the supernatants were evaporated to dryness using a speedvac cold trap concentrator. Each dried sample was reconstituted in a 50 μl methanol: toluene (9:1, v/v) mixture, vortexed for 10s, transferred to glass vials with micro-inserts, capped immediately, and injected into the Ultra High-Performance Liquid Chromatography (UHPLC)-MS/MS system in multiple reaction monitoring (MRM) mode. The oleamide identification was confirmed by comparing the retention time and the qualifier/quantifier ion of the analytes in the sample with those obtained from the authentic analytical standard. The calibration curve was employed for the quantitative determination of oleamide in the samples (see supplementary figure 2 and MSMS parameters Supplementary Table 3). For details on LC-MS/MS oleamide quantitation with the pure deuterated standard see supplementary methods.

## Abbreviations

AD: Alzheimer’s Disease
MCI: mild cognitive impairment
cMCI: MCI converted to Alzheimer’s
sMCI: MCI stable
MMSE: mini mental state examination
NC: normal cognition
ML: machine learning
LR: logistic regression
MLP: multi-layer perceptron network
RF: random forest
SVM: support vector machine.

## Code availability

The code used to preprocess the EMIF-AD proteomic and metabolomic data as well as the code to create the machine learning models and represent the results are available on GitHub (https://github.com/aliciagp/ML-multiomics).

## Data availability

The generated metabolomic and proteomic data in EMIF is considered sensitive patient data and can therefore not be publicly available in compliance with the European privacy regulations governed by GDPR and according to limitations included in the informed consents signed by the study participants. Data are available upon request by contacting the EMIF-AD data hub steering committee via the academic EMIF-AD lead, Prof. Pieter Jelle Visser, and data access coordinator Dr. Stephanie Vos. Requests should include the following information, name and contact details of the person requesting the data, aims and objectives, study design and methods, requested molecular data and clinical variables, outcomes, timelines and cohorts of interest in the EMIF-AD catalogue (https://emif-catalogue.eu/login). Requests will be subject to consideration by the steering committees of the cohorts and the management board. Time frame for a response will be within 4 months. Data requests under agreement will be subject to appropriate confidentiality obligations and restrictions.

The averaged protein and metabolites plasma levels for each diagnosis group (NL, sMCI, cMCI and NL) have been deposited on GitHub (https://github.com/aliciagp/ML-multiomics).

## Supplementary material

### Supplementary methods. Analysis of oleamide by UHPLC-MS/MS

Samples were analyzed in dynamic multiple reaction mode (dMRM) on an Agilent 1290 Infinity UHPLC system connected to an Agilent 6460 triple quadrupole (QqQ) mass spectrometer (MS) from Agilent Technologies Inc. (Santa Clara, CA, USA). Waters HSS T3 2.1 × 100 mm, 1.8 μm column protected by a C18 HSS T3 VanGuard Pre-column (100Å, 1.8 µm, 2.1 mm × 5 mm) both from (Waters, Taastrup, DK.). The column temperature was maintained at 45°C throughout the run with a flow rate of 0.4 mL/min. The injection volume was 5 μL, and a binary solvent mixture was used. Solvent A contained water and solvent B acetonitrile/isopropanol (67/33) (v/v). 0.1% formic acid (v/v) was added to both solvents. The following gradient was used for positive mode analysis: 0-1 min, 60% A, 1-2 min, ramping to 20% A, 2-8 min, 0% A, 8-9 min, 0% A, 9-9.2 min, ramping back to 60% A, 9.2-12 min, 60% A. Instrument-dependent parameters for mass spectrometry were as follows: The nitrogen drying gas flow and the temperature was 12 L/min and 325 °C, respectively. The capillary voltage was 3500. The nebulizer pressure was controlled at 45 psi. The nitrogen sheath gas flow and temperature were kept at 11.0 L/min and 325 °C, respectively. Supplementary Table 6 summarizes dMRM transitions, retention times, fragmentator voltages, and collision energies used for oleamide.

**Supplementary figure 1.**
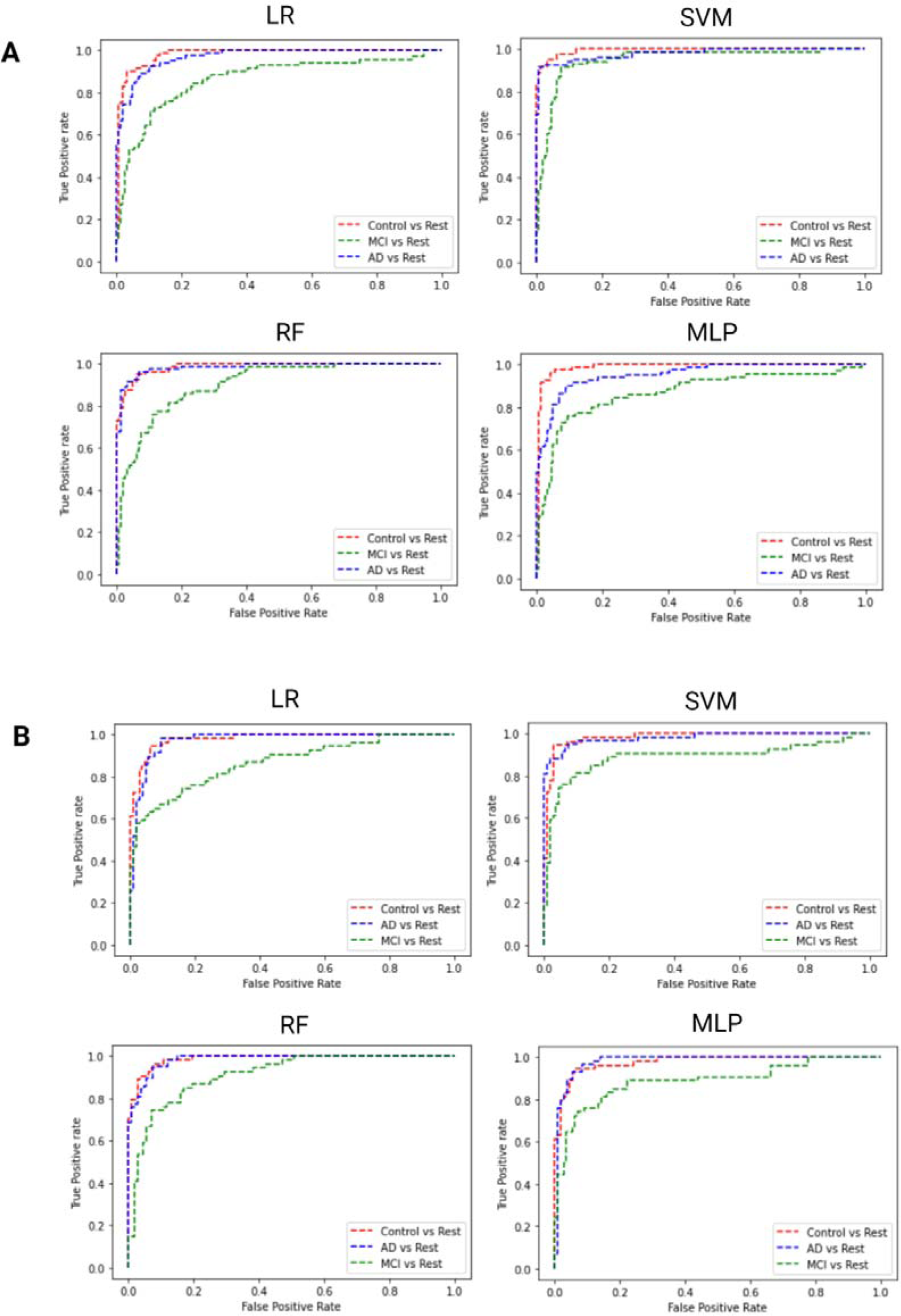
Multiclass models ROC curves. Models performance to classify NC, MCI and AD donors using four different machine learning algorithms using (**A)** proteins and clinical features, **(B)** metabolites and clinical features.

**Supplementary figure 2.**
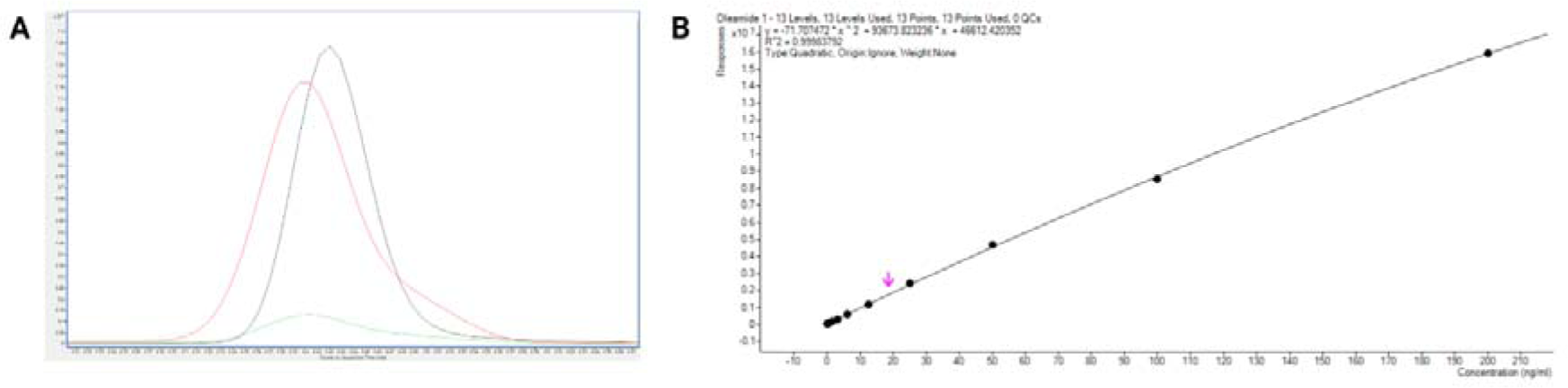
Identification and quantification of oleamide. **(A)** The chromatogram obtained through LC-MS/MS with multiple reaction monitoring (MRM) mode confirmed the presence of oleamide in both the extracellular vesicles (EVs) isolated from the plasma of the AD sample (red) and the plasma of the control samples (green). The identification of oleamide was confirmed by comparing the retention time and the qualifier/quantifier ion ratio (with a tolerance of 20%) of the oleamide in the sample with those obtained from the authentic analytical standard (black). **(B)** The calibration curve was employed for the quantitative determination of oleamide in the samples.

**Supplementary Table 1. Top 20 features selected per algorithm for multiclass models (proteins and metabolites separately) and MCI conversion models (both proteins and metabolites).** See excel file.

**Supplementary Table 2.** Performance of the MCI conversion models for proteins and metabolites for each algorithm. Standard errors indicated in brackets represent cross-validation error.

| Omics data | Algorithm | ROC<br>Training data | Sensitivity<br>Training data | Specificity<br>Training data |
| --- | --- | --- | --- | --- |
| Clinical data,<br>proteins<br>and metabolites | LR | 0.640 (0.048) | 0.614 (0.091) | 0.582 (0.089) |
|  | SVM | 0.630 (0.043) | 0.620 (0.090) | 0.575 (0.101) |
|  | RF | 0.662 (0.067) | 0.621 (0.112) | 0.588 (0.092) |
|  | MLP | 0.641 (0.058) | 0.633 (0.103) | 0.549 (0.091) |

**Supplementary table 3.** Genetics, transcriptomes and proteomes studies where the candidate proteins were also associated with an AD-related phenotype. We checked whether changes in levels of selected proteins are relevant in AD brain at genetics, transcriptomics and proteomic level (https://agora.adknowledgeportal.org/). Brain eQTL indicates whether or not this gene locus has a significant expression Quantitative Trait Locus (eQTL) based on an AMP-AD consortium study. RNA/protein expression change in AD brain indicates whether or not this gene shows significant differential (protein) expression in at least one brain region based on AMP-AD consortium work. In addition, we also checked if blood level of candidate proteins were already associated with AD-related phenotype (Kiddle, S. J. et al 2014).

| UniProt ID | Protein name | Gene symbol | Brain eQTL | RNA expression change in AD brain | Protein expression change in AD brain | Blood level of protein change in AD |
| --- | --- | --- | --- | --- | --- | --- |
| P43155 | Carnitine O-acetyltransferase | CRAT | ✓ | ✓ |  |  |
| P37840 | Alpha-synuclein | SNCA | ✓ | ✓ | ✓ |  |
| Q8WXH2 | Junctophilin-3 | JPH3 | ✓ | ✓ |  |  |
| P27918 | Properdin | CFP |  | ✓ | No data |  |
| O43692 | Peptidase inhibitor 15 | PI15 | ✓ | ✓ | No data |  |
| P01298 | Pancreatic hormone | PPY |  | No data | No data | ✓ |
| P04054 | Phospholipase A2 | PLA2G1B |  | No data | No data |  |
| Q9NS68 | Tumor necrosis factor receptor superfamily member 19 | TNFRSF19 | ✓ | ✓ | No data |  |

**Supplementary table 4.**
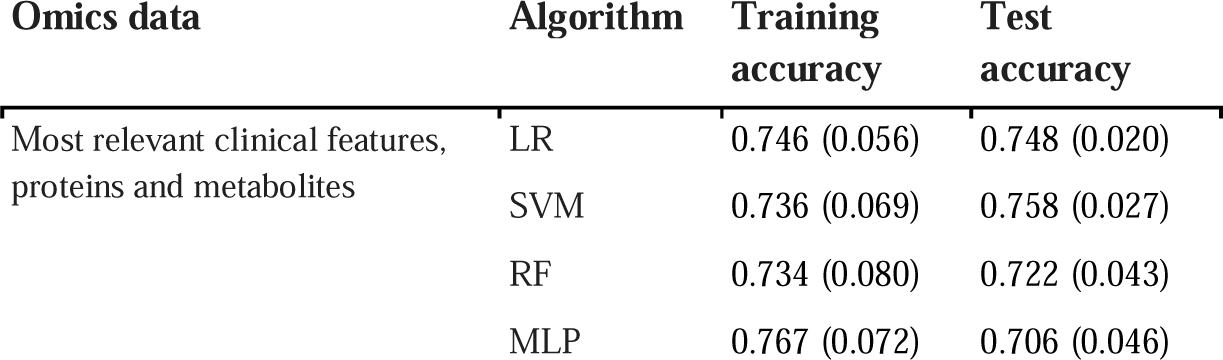
MCI conversion models performance for each algorithm. Standard errors indicated in brackets represent nested cross-validation error.

**Supplementary Table 5.** Demographics of participants included in the MCI conversion approach by diagnosis. One-way analysis of variance (ANOVA) and chi-square tests were used to compare continuous and binary variables, respectively. Aβ status was defined by the CSF Aβ42/40 of the central analyses, using a cutoff value of < 0.061 to determine abnormality. Standard errors are indicated in the brackets.

| Characteristics | sMCI | cMCI | *P*<br>value |
| --- | --- | --- | --- |
| n | 103 | 91 | NA |
| Age mean (SD) y | 69.17 (8.61) | 70.76 (7.49) | 0.176 |
| Male sex N (%) | 52 (50.5) | 50 (54.9) | 0.634 |
| Education mean (SD) y | 11.35 (3.24) | 11.25 (3.45) | 0.832 |
| MMSE mean (SD) | 26.15 (2.69) | 24.98 (2.82) | 0.003 |
| Phosphorylated TAU mean (SD) | 66.12 (33.58) | 83.91 (37.00) | 0.001 |
| Amyloid positive status (%) | 71 (68.9) | 81 (89.0) | 0.001 |
| Priority Attention Z-score mean (SD) | -0.64 (1.40) | -0.96 (1.52) | 0.121 |
| Priority Language Z-score mean (SD) | -0.76 (1.27) | -1.30 (1.37) | 0.004 |
| Priority Memory Immediate Z-score mean (SD) | -1.26 (1.32) | -1.78 (1.18) | 0.004 |
| APOE E4+ N (%) | 55 (53.4) | 58 (63.7) | 0.190 |

**Supplementary Table 6.** MS/MS parameters for the analysis of primary fatty acid amides by dMRM in positive mode. Q1, quadrupole 1 m/z ratio; Q3, quadrupole 3 m/z ratio; R_t,_ retention time (min); CE, collision energy.

| Analyte | Q1 (m/z) | Q3 (m/z) | R<sub>t</sub> (min) | Frag (v) | CE (v) |
| --- | --- | --- | --- | --- | --- |
| Oleamide | 282.2 | 69.2 | 4.6 | 92 | 21 |
|  |  | 55.2 |  | 92 | 45 |

